# The Morges Strategy for Yaws Eradication: the First Largescale Total Community Treatment with Azithromycin Against Yaws in the Congo-Basin, using a Novel Model

**DOI:** 10.1101/2024.11.07.24316738

**Authors:** Earnest Njih Tabah, Alphonse Um Boock, Chefor Alain Djam, Gilius Axel Aloumba, Boua Bernard, Nzoyem Colin Tsago, Irine Ngani Nformi, Loic Douanla Pagning, Elisaberth Baran-A-Bidias, Christian Elvis Kouayep-Watat, Smith Afanji, Ebai George, Marielle Patty Ngassa, Bonaventure Savadogo, Serges Tchatchouang, Valerie Donkeng, Yves Thierry Barogui, Sara Eyangoh, Kingsley Bampoe Asiedu

**Affiliations:** National Yaws, Leishmaniasis, Leprosy and Buruli ulcer Control Programme, Ministry of Public Health, Yaounde, Cameroon; Department of Public Health, Faculty of Medicine and Pharmaceutical Sciences, University of Dschang, West Cameroon; Research Initiative in Tropical and Community Health, Yaounde, Cameroon; FAIRMED, Health for the Poorest, Bern, Switzerland; National Neglected Tropical Diseases Control Programme, Ministry of Health, Brazzaville, Congo; National Neglected Tropical Diseases Control Programme, Ministry of Health and the Population, Bangui, Central African Republic; OCEAC, Yaounde, Cameroon; Centre Pasteur du Cameroun, Yaounde, Cameroon; WHO Regional Office for Africa, Brazzaville, Congo; Department of Control of Neglected Tropical Diseases, WHO, Geneva, Switzerland

**Author notes:** **Corresponding Author** Earnest Njih Tabah.

**Keywords:** Novel model, TCT, yaws, prevalence, Congo-Basin, CEMAC

## Abstract

**Context and Justification:** Yaws is targeted for eradication by 2030. Total Community Treatment with azithromycin (TCT), a major component of the eradication strategy, has witnessed only three pilots since 2012. We implemented the first large-scale TCT in the Congo-Basin of Central Africa using a novel model.

**Methodology:** We implemented a novel 3-phase TCT model in 17 health districts of the Congo-Basin, spanning 3 countries. Two rounds were implemented in Cameroon, and one round each in Central African Republic (CAR) and the Republic of Congo; targeting 1,530,014 people (144,934(9.5%) Pygmies and 1,304,410(90.5%) Bantus). TCT was followed by post-campaign active surveillance, treatment of yaws cases and their contacts.

**Results:** All 17 health districts were confirmed for yaws endemicity. Overall, 1,456,691 (95.21%; 95%CI: 95.17%-95.24%) persons were treated in the first round of TCT, including 552,356/594411 (92.92%; 95%CI: 92.86%-92.99%) in Cameroon, 359,810/373,994 (96.21%; 95%CI: 96.15%-96.27%) in CAR, and 544,526/561,609 (96.96%; 95%CI: 96.91%-97.00%) in Congo. For the second round implemented only in Cameroon, 615,503/642,947 (95.73%; 95%CI: 95.68%-95.78%) were treated. There was a 3-percentage-point increase in therapeutic coverage between the first and second round (P-value<0.001), and from 89.2% to 93.1% (P-value<0.001) among the Pygmies. The prevalence of active yaws decreased from 6.5% to 0.4% (P-value<0.001) overall; and from 6.31% to 0.23% (P-value<0.001, OR=28.67, 95%CI (20.86-40.09) in Cameroon, from 2.4% to 0.8% (P-value=0.002, OR=3.19, 95% CI: 1.45-8.00) in CAR, and from 10.8% to 3.7% (P-value<0.001, OR=3.18, 95%CI: 2.09 – 5.01) in Congo.

**Conclusion:** A novel TCT model was successfully implemented at largescale in the Congo-Basin, achieving above recommended threshold of therapeutic coverages. The prevalence of active yaws dropped remarkably following the TCT, however complete interruption of yaws transmission was not achieved. In recommending the novel model of TCT to endemic countries, we suggest that at least three rounds, at six-monthly intervals, are implemented for complete interruption of yaws transmission.

**Author summary:** Yaws, a skin-NTD that affects mainly children resurfaced among the Pygmies of the Congo-Basin around 2007 after eradication in the late 1970s. Renewed efforts for eradication guided by the Morges strategy are being promoted by the WHO. In this regard, we a novel model for Total Community treatment (TCT) with azithromycin in 17 health districts of the Congo-Basin spanning three countries including Cameroon, CAR and The Republic of Congo. This was the largest scale so far, since the inception of the Morges strategy. Of a total population of 1,530,014 targeted in the first round of TCT, 1,456,691(95.21%) persons were treated including 552,356 in Cameroon, 359,810 in CAR, and 544,526 in Congo. For the second implemented only in Cameroon, 615,503 (95.73%) out of 642,947 people targeted were treated. Our novel model permitted the achievement of therapeutic coverages surpassing the 90% threshold recommended by the WHO. The TCT led to a remarkable reduction of the prevalence of active yaws in the zone, but however, did not interrupt its transmission. In recommending the novel model of TCT to yaws endemic countries, we think that at least three rounds at six-monthly intervals are necessary for complete interruption of yaws transmission.

## 1. **I**ntroduction

Yaws, one of the endemic treponematoses, is a Neglected Tropical Disease (NTD), caused by *Treponema pallidum* subspecies *pertenue*(1). It affects persons of both genders and all ages but predominantly children below 15 years of age(2), leading to primary and secondary skin lesions that are highly contagious, and tertiary non-infectious but destructive lesions of the cartilage and the bones(1–3). Transmission is by direct non-venereal human-to-human contact, via exudate from infectious lesions and spread is facilitated by overcrowding and poor personal and environmental hygienic conditions(3). Yaws exhibits clinical polymorphism, with over seven clinical forms: papilloma, papule, nodule, ulcers, macular, squamous-macular, plantar/palmar and osteo-periostitis(1,4), posing a challenge for clinical diagnosis. A two stage serological rapid diagnostic testing for screening and confirmation of yaws is widely recommended (4–7).

Yaws was a serious public health problem in the 1950s involving over 46 tropical countries worldwide(1). A joint eradication effort led by WHO and the United Nations Children’s Fund (UNICEF) through mass treatment campaigns with single-dose benzathine penicillin injection, resulted in the treatment of over 300 million people between 1952 and 1964(8). At the end of these campaigns, the burden of yaws was reduced by 95% globally, corresponding to an absolute reduction in prevalence from 50 million to 2,5million cases(1,8,9).

After this achievement, yaws surveillance efforts were relented, leading to its resurgence in the late 1970s in parts of west and central Africa, Asia and the pacific(10–14). Following this resurgence, the 31^st^ World Health Assembly adopted a new resolution: WHA31.58, calling for renewed efforts by endemic countries, especially those with resurgence, to develop integrated control programmes for yaws eradication, with emphasis on surveillance(15). However, this resolution was not well implemented in many affected countries due to lack of political will, funding issues, competing health priorities, and weak health systems(16). Consequently, by 2012, as many as twelve previously endemic countries including Cameroon, CAR, and The Republic of Congo, were actively reporting yaws(8,17).

Meanwhile, there were also very important developments that brought about new hope and momentum for a renewed yaws eradication effort:

− A successful clinical trial in the Papua New Guinea in 2012 proved azithromycin oral tablets to be as efficacious as benzathine penicillin in the treatment of yaws(18).
− India, the second most populated country in the world was certified yaws-free by WHO in 2016 (8,19).
− New point-of-care tests for yaws screening and confirmation as well as PCR technology for monitoring of yaws resistance were now available(4).
− Yaws was targeted for eradication by 2020 in the first WHO NTDs Roadmap for the period 2012-2020(20).
− The International Task Force for Disease eradication, after critically examining these developments, and the feasibility, lent its support to the renewed yaws eradication momentum(8,21).

To operationalize the yaws eradication efforts, a WHO expert consultative meeting held in Morges, Switzerland in 2012 and developed a strategy for yaws eradication, code named “The Morges Strategy”(22–24). The eradication strategy was subsequently accompanied by a guideline for programme managers(25). Box 1. Shows the four components

### Box 1. Components of the yaws eradication strategy

**Total community treatment (TCT):** Entire endemic community receives treatment initially, irrespective of the number of active clinical cases or the prevalence of yaws disease. Two or three rounds of TCT at 6monthly intervals, and achieving a population coverage of > 90%, is essential to interrupt transmission or significantly reduce prevalence of cases.

**Total targeted treatment (TTT):** All new yaws cases and their contacts (household, frequent family friends, schoolmates and playmates) are treated after the initial or in-between rounds of TCT campaigns.

**Health system approach:** Resurveys for case-finding, treatment and surveillance are continued by health system personnel. The health system would be responsible for:

i. confirming and treating suspected as well as missed cases including their contacts, and
ii. (ii) sustaining public awareness and interest through health promotion, case-finding, and treatment activities.

**Essential supportive measures:** The central and district yaws eradication teams should provide technical and logistic support to health facilities throughout the yaws eradication process.

of the yaws eradication strategy(22,25).

After the publication of the Morges strategy, pilot implementation were carried out in the Lihir Island, Papua New Guinea where 13,490(83.8%) of 16,092 population(26) and in the Abamkron sub-district in Ghana where 14,548 (89%) of 16,287 population(27) were treated with single dose azithromycin respectively. These pilots were limited in scope.

In this paper, we report the implementation and outcomes of the first large-scale total community treatment with azithromycin (TCT) against yaws, in 17 health districts of the Congo Basin, spanning three countries of the CEMAC subregion and covering a total population of 1,530,014, between 2020 to 2023, using a novel model.

## Methods

### 1.1. Site, Setting and Population

We conducted an intervention to demonstrate the feasibility of TCT at a large scale in 17 health districts located in the Congo-Basin, spanning three countries of the CEMAC subregion namely Cameroon, The Central African Republic (CAR) and The Republic of Congo. These included ten districts in Cameroon: namely Abong Mbang, Doumé, Lomie, Mbang, Nguelemendouka, Messamena, Moloundou, Ndélélé, Yokadouma in the east region, and Djoum in the south region; four health districts in the Republic of Congo namely Ouesso, Sembe-Souanke in Sangha department and Impfondo and Dongou-Enyelle in Likouala department; and three health districts in CAR including Mbaiki, Boda and Sangha-Mbaere (Fig. 1).

**Fig 1:**
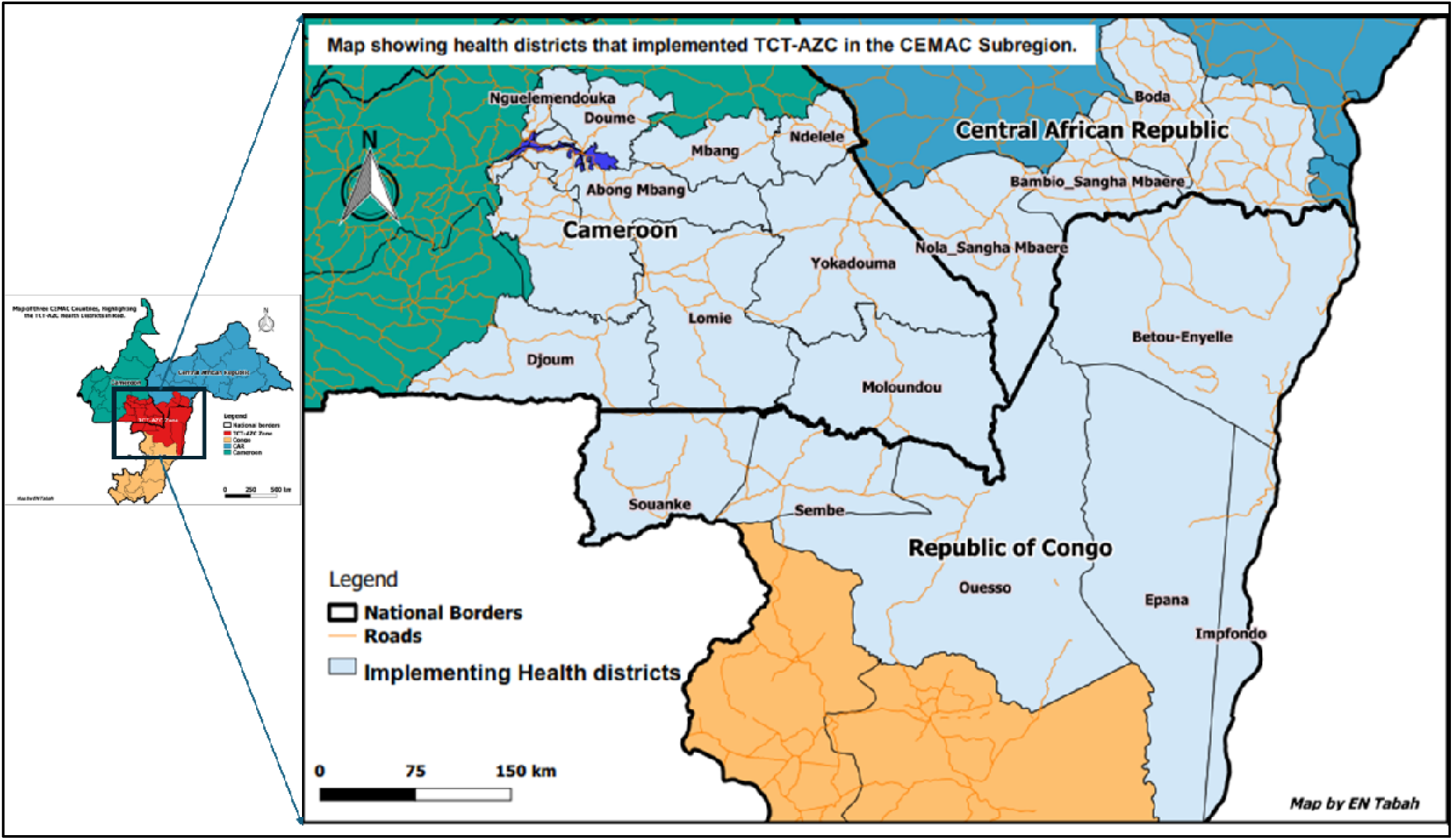
Map highlighting the 17 health districts of the Congo Basin spanning three countries (Cameroon, CAR, and Congo.), that implemented the TCT. The Congo-Basin is home to the indigenous Pygmy population.

The zone is covered by a dense tropical rainforest and have many swamps and rivers which are loaded with a rich and diversified wildlife(28,29). The climate is warm and humid with temperatures ranging from 22°C-30°C. It has two seasons: the rainy season that runs from March-November, and the dry season from December-February. The average annual rainfall ranges from 1500mm to 1800mm but can go to a maximum over 10,000mm in the heart of the rainy season(30). The zone is enclave and most of the villages and pygmy camps are accessed only through foot paths. The roads when they exist, are passable only in the dry season. This is compounded in some places by large rivers and streams without bridges. The area is also poorly covered by portable water as well as telephone and electricity networks.

The zone was inhabited by 1,530,014 people in the mid of 2022 (31–33) with Cameroon accounting for 594,411(38.85%), the Central African Republic 561,609 (36.71%) and the Republic of Congo 373,994 (24.44%) respectively. The population consists of about 1,304,410 (90.53%) Bantus and 144,934 (9.47%) indigenous Pygmies (see Table 1). The whole of this population was targeted for the TCT, with a particular emphasis on the indigenous Pygmy, populations who are most vulnerable to yaws.

**Table 1.** Distribution of population of the targeted health districts by race and by age.

| Health District /<br>Country | Total pop.<br>Targeted | Distribution by race |  | Distribution by age |  |  |
| --- | --- | --- | --- | --- | --- | --- |
|  |  | Baka<br>pop. | Bantu<br>pop. | 0-59<br>months | 6-15 yrs | 16+yrs |
| Cameroon |  |  |  |  |  |  |
| Abong-Mbang | 86,089 | 5,405 | 80,684 | 12,827 | 23,244 | 50,018 |
| Doume | 53,094 | 1,990 | 51,104 | 7,911 | 14,335 | 30,848 |
| Lomie | 37,668 | 12,754 | 24,914 | 5,613 | 10,170 | 21,885 |
| Mbang | 34,274 | 5,336 | 28,938 | 5,107 | 9,254 | 19,913 |
| Messamena | 30,023 | 2,924 | 27,099 | 4,473 | 8,106 | 17,443 |
| Moloundou | 48,077 | 6,056 | 42,021 | 7,163 | 12,981 | 27,933 |
| Ndelele | 101,680 | 2,677 | 99,003 | 15,150 | 27,454 | 59,076 |
| Nguelemendouka | 39,186 | - | 39,186 | 5,839 | 10,580 | 22,767 |
| Yokadouma | 118,598 | 15,762 | 102,836 | 17,671 | 32,021 | 68,905 |
| Djoum | 45,722 | 4,810 | 40,912 | 6,813 | 12,345 | 26,564 |
| Total Cameroon | 594,411 | 57,714 | 536,697 | 88,567 | 160,491 | 345,353 |
| Republic of Congo |  |  |  |  |  |  |
| Impfondo | 130,625 | 13,063 | 117,563 | 19,463 | 35,269 | 75,893 |
| Enyelle Betou | 109,643 | 10,964 | 98,679 | 16,337 | 29,604 | 63,703 |
| Ouessou | 96,525 | 9,653 | 86,873 | 14,382 | 26,062 | 56,081 |
| Sembe | 37,201 | 3,720 | 33,481 | 5,543 | 10,044 | 21,614 |
| Total Congo | 373,994 | 37,399 | 336,595 | 55,725 | 100,978 | 217,291 |
| Central African<br>Republic (CAR) |  |  |  |  |  |  |
| Boda | 122,516 | 12,252 | 110,264 | 18,255 | 33,079 | 71,182 |
| Mbaiki | 289,117 | 20,845 | 187,602 | 31,059 | 56,281 | 121,108 |
| Sangha Mbaere | 149,976 | 14,998 | 134,978 | 22,346 | 40,494 | 87,136 |
| Total CAR | 561,609 | 48,094 | 432,845 | 71,660 | 129,854 | 279,426 |
| Grand Total | 1,530,014 | 144,934 | 1,304,410 | 215,952 | 391,323 | 842,069 |

The Pygmies are indigens of the Congo-Basin rainforest where they have lived for millions of years(29,34–36). Their mode of life as well as their low population density, render them very vulnerable. Their lifestyle is organized around the forest and its resources, where they live in a semi-nomadic mode, and depend on it for hunting and gathering of non-timber resources for their basic needs(35–37). The Pygmies live in small groups within which members of a whole family all live in a small hut constructed from leaves and sticks(34). The overcrowding, promiscuity, and little or no attention to personal hygiene in these groups favour the transmission of diseases(13,38) and have been attributed to the re-emergence of yaws(39). However, limited access to conventional health care, and probable indigenous knowledge has led to the development of a reputable indigenous health care system, predominantly based on medicinal plants(38)

### 1.2. Confirmation of endemicity status of the targeted health districts

Prior to the implementation of the Morges Strategy for yaws eradication, the World Health Organisation (WHO) requires that yaws endemicity status of the implementation units (health districts in this case) is confirmed. 0

In Cameroon, verification of yaws endemicity status in the ten targeted health districts was conducted in three phases: The investigation and riposte of an epidemic outbreak that occurred in Lomie health district in May 2017 offered the first opportunity. This outbreak was the fourth in the Lomie health district following the ones in 2007, 2008 and 2010(40), respectively. These outbreaks each time sparked off in a Baka pygmy camp before spreading to the rest of the Baka camps and Bantu settlements. During the investigation of this fourth outbreak in May 2017, 471(25%) clinical cases of yaws were detected among 1885 persons examined, and 154 (8.17%) of them were serological confirmed with the DPP syphilis screen and confirm kit(41) (Table 1). A second survey was organized in September 2017, in four health districts sharing borders with the Lomie health district, including Abong-Mbang, Djoum, Moloundou and Yokadouma, which also harbour the Baka pygmy population. Each of these four districts were serologically confirmed yaws endemic with prevalence varying from 0.92% in Djoum to 7.32% in Moloundou. The third survey was conducted in March 2020 in the four health districts including Mbang, Ndelele, Doume, Nguelemenendouka and Messamena completed the queue. The prevalence of active yaws was found to range from 2.65% in Nguelemendouka to 12.7% in Mbang health districts (Table 2).

**Table 2:** Prevalence of yaws in the surveyed health districts before TCT.

| Event | Health District | N° of persons Examined | Clinical yaws |  | DPP confirmed (active yaws) |  |
| --- | --- | --- | --- | --- | --- | --- |
|  |  |  | n | % | n' | % |
| **Cameroon** | **Total** | **3,584** | **1,072** | **30%** | **226** | **6.31%** |
| Investigation of out-break of May 2017 | Lomie | 1,885 | 471 | 25% | 154 | 8.17% |
| Survey of August 2017 | Moloundou | 287 | 116 | 40% | 21 | 7.32% |
|  | Yokadouma | 864 | 266 | 31% | 30 | 3.47% |
|  | Abong_Mbang | 92 | 91 | 99% | 2 | 2.17% |
|  | Djoum | 109 | 7 | 6% | 1 | 0.92% |
| Survey of March 2020 | Mbang | 63 | 22 | 35% | 8 | 12.70% |
|  | Ndelélé | 45 | 19 | 42% | 3 | 6.67% |
|  | Doumé | 54 | 26 | 48% | 2 | 3.70% |
|  | Nguelemendouka | 113 | 37 | 33% | 3 | 2.65% |
|  | Messamena | 72 | 17 | 24% | 2 | 2.78% |
| **Republic of Congo** | **Total** | **1,627** | **987** | **60,66%** | **176** | **17,83%** |
| Survey of February 2021 | Ouessou | 417 | 189 | 45,32% | 32 | 16.9% |
|  | Sembe - Souanke | 392 | 207 | 52,80% | 16 | 7,7% |
|  | Impfondo | 316 | 213 | 67,40% | 46 | 21,15% |
|  | Enyelle – Betou | 502 | 378 | 75,29% | 82 | 21,6% |
| **Central African Republic (CAR)** | **Total** | **1,469** | **477** | **32,47** | **35** | **7,33%** |
| Survey of March 2021 | Boda | 302 | 72 | 23,84% | 6 | 8,33% |
|  | Mbaïki | 665 | 207 | 31,13% | 10 | 4,83% |
|  | Sangha Mbaéré | 502 | 198 | 39,44% | 19 | 9,59% |

In the Republic of Congo, yaws endemicity survey was conducted in February 2021 in four health districts namely: Ouesso, Sembe-Souanke, Impfondo and Enyelle-Betou, where active yaws prevalence was found to range from 7.7% in Sembe-Souanke to 21.6% in Enzelle-Betou (Table 2).

The yaws endemicity survey was conducted in three health districts (Mbaiki, Boda and Sangha-Mbaere) of the Republic of Central Africa in March 2021, where the prevalence of active yaws ranged from 4.83% in Mbaiki to 9.59% in Sangha-Mbaere (Table 2).

### 1.3. Development of a novel TCT campaign model

For reasons of paucity in experiences on TCT against yaws and the need to build a model adapted to the Congo Basin context, experiences from other national programs versed with MDAs and other community-based interventions were sought. The national onchocerciacis program (PNLO) experienced with ivermectin MDA, the EPI program experienced with national immunisation days, and the malaria program experienced with community distribution of mosquito nets, were brought together to share their experiences with the National Yaws, Leishmaniasis, Leprosy and Buruli ulcer Control Programme in Cameroon.

The various experiences were capitalized and served as a basis for the development of the model for the TCT against yaws in the Congo Basin. Briefly, the model developed to be fully implemented within a period of 15 to 21 days, included three main phases namely planning, execution and evaluation of the TCT campaign. Fig. 2 show details of the process activities and their key outcomes of the model that was developed. This model was successfully implemented in two rounds of TCT in Cameroon and one round each in the Congo and in RCA.

**Fig. 2.**
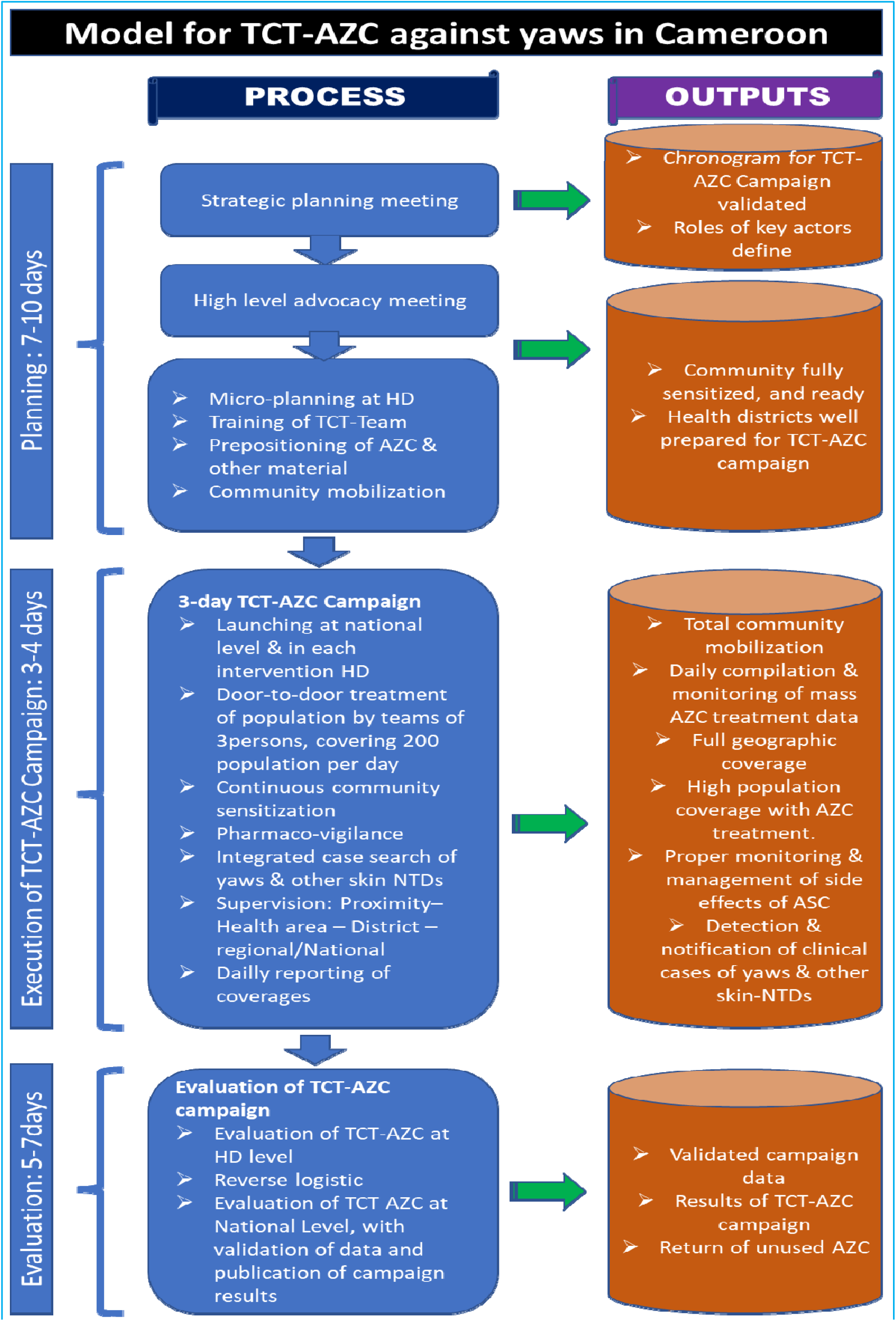
Novel Model for TCT against yaws developed and implemented in Cameroon, CAR and Congo. (TCT=Total Community Treatment, AZC= Azithromycin, HD= Health District)

### 1.4. The Planning phase

The planning phase included: a strategic planning meeting, high-level advocacy, and health district level micro-planning.

#### 1.4.1. The strategic planning meeting

The strategic planning meeting presided over by the National Programme Manager in charge of skin NTDs, brought together national level stakeholders from the national programme; the Regional Delegation of Public Health and the District Health Management Teams (DHMTs) from the targeted health districts; the WHO country office; and Support Partners. During this strategic planning meeting, the various actors involved in execution of the TCT campaign were identified and their roles precisely defined (see box 2).

##### Box 2. Key actors of TCT Campaign and their roles

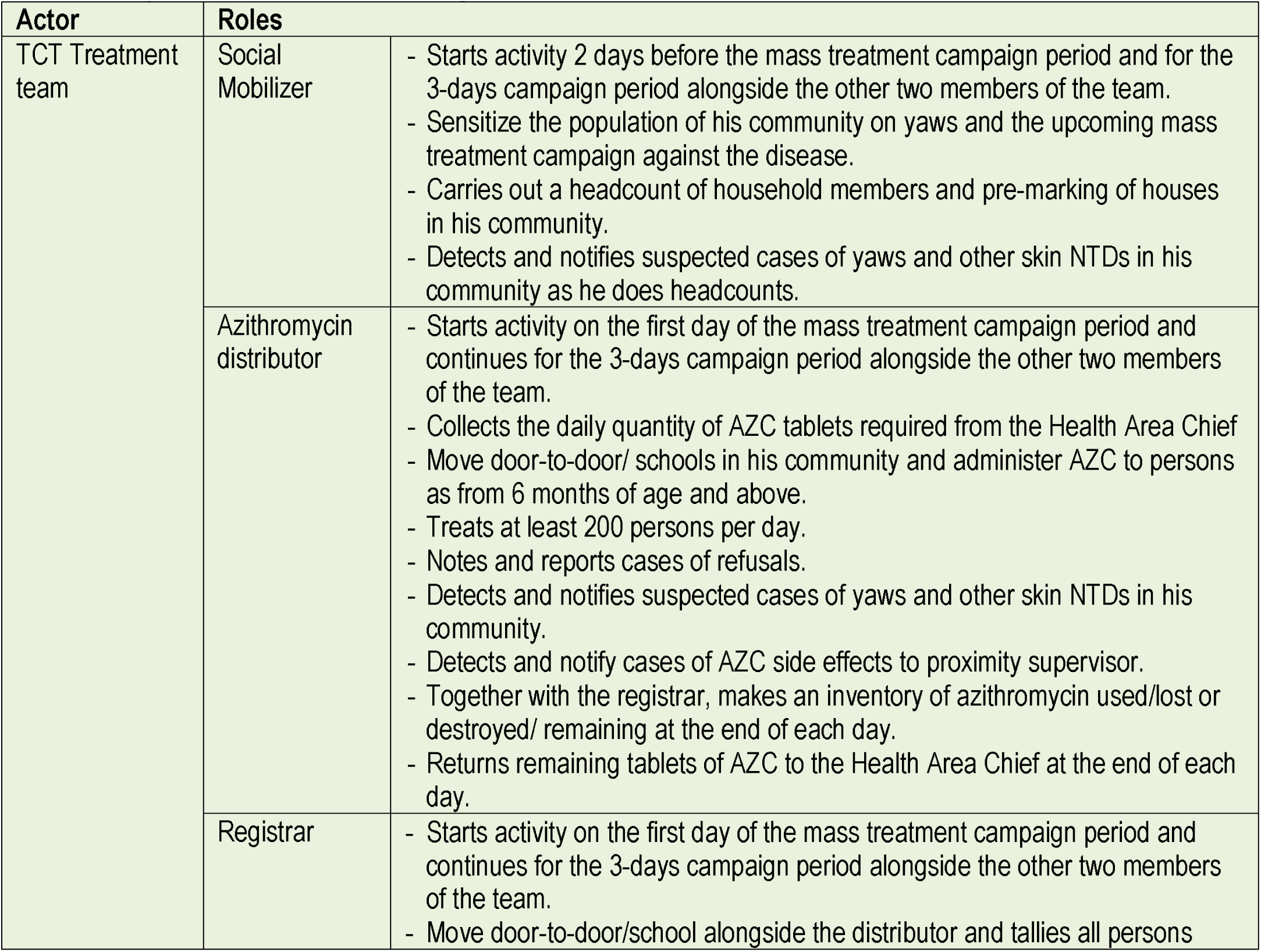
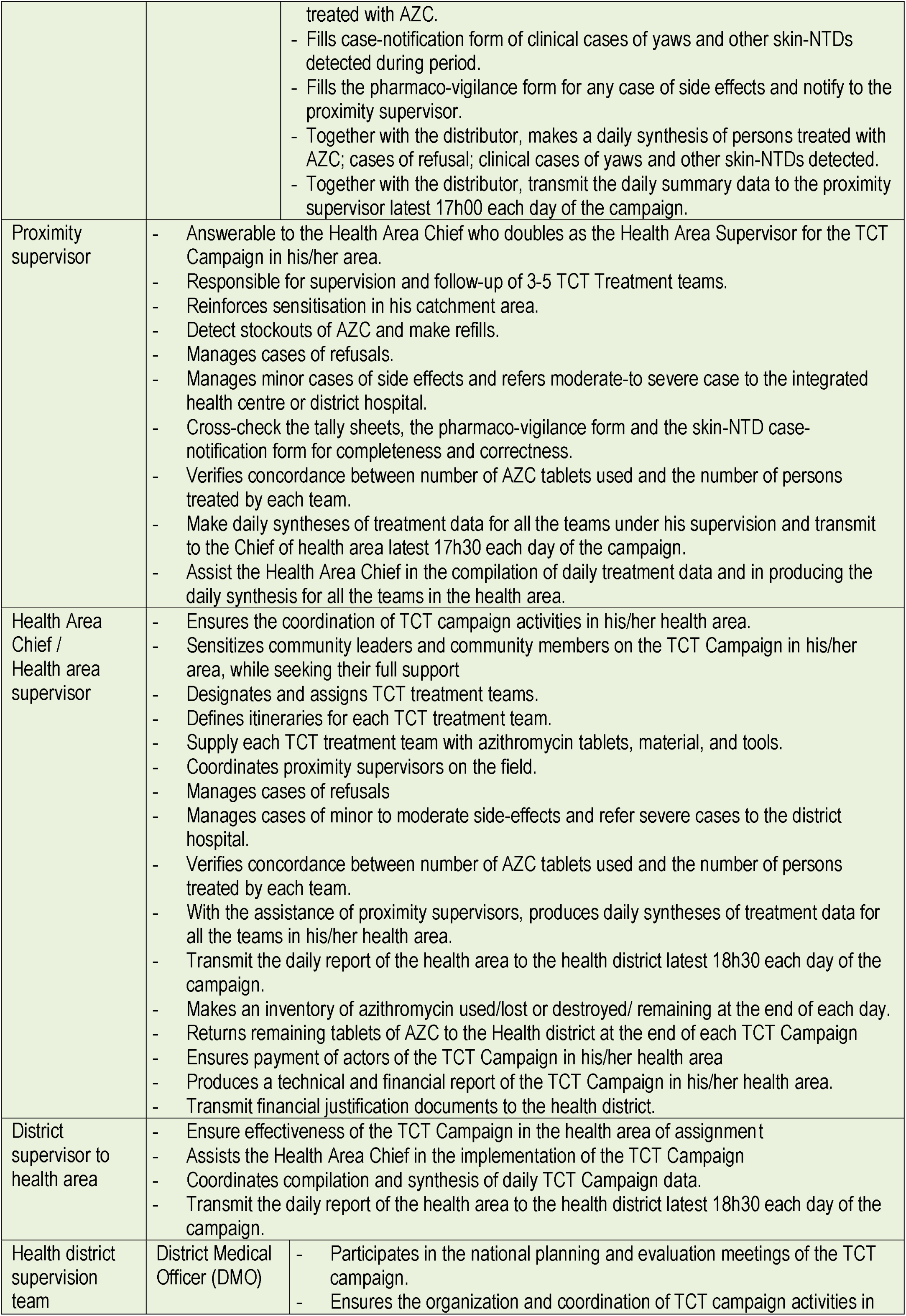
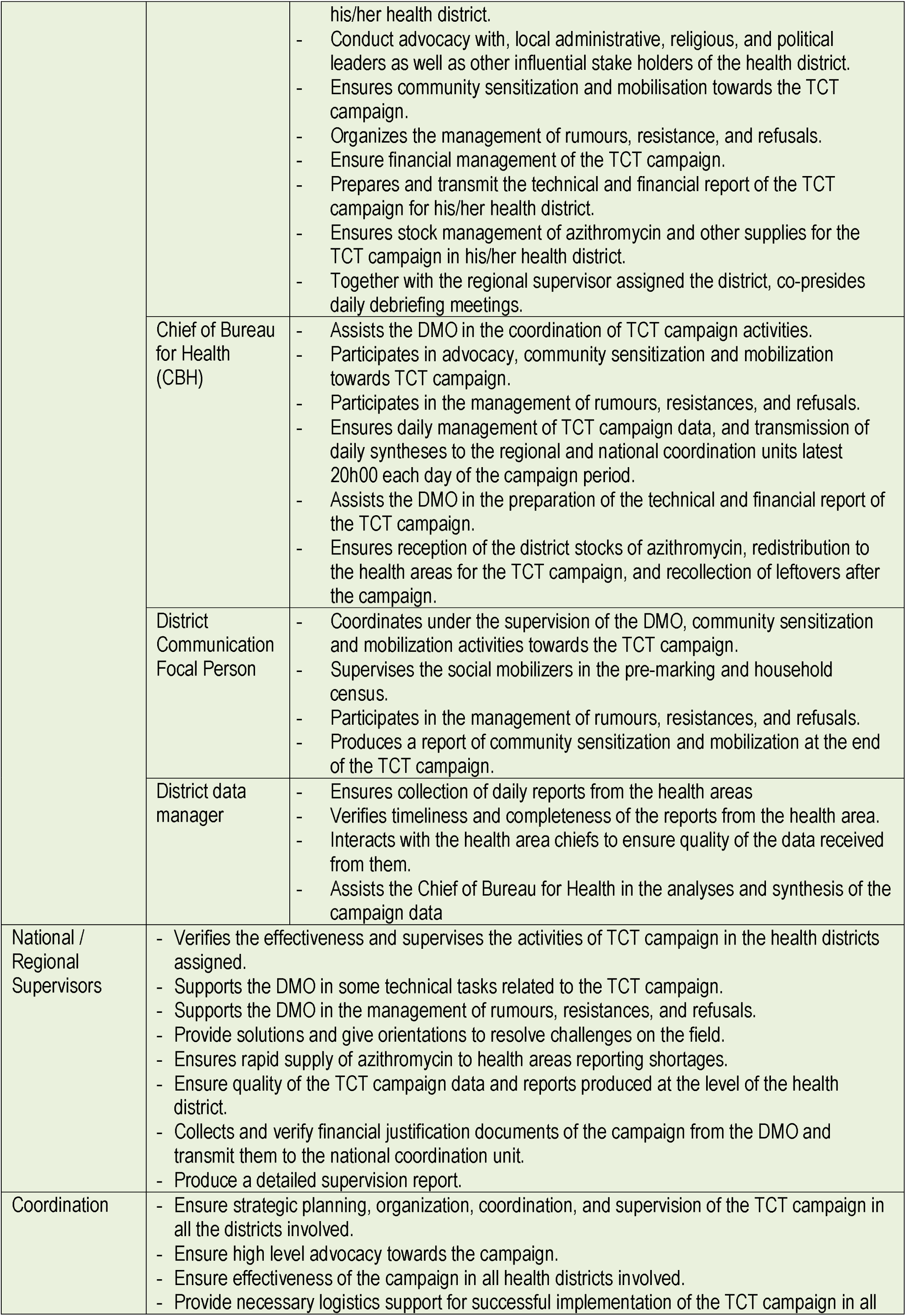
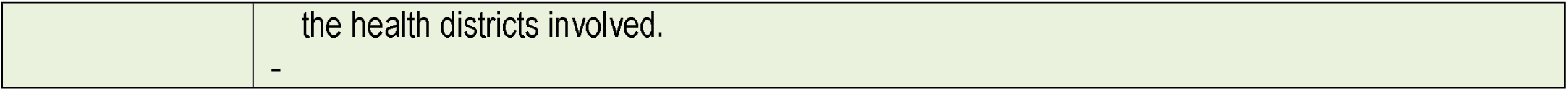

| Actor | Roles |  |
| --- | --- | --- |
| TCT Treatment team | Social Mobilizer | - Starts activity 2 days before the mass treatment campaign period and for the 3-days campaign period alongside the other two members of the team.<br>- Sensitize the population of his community on yaws and the upcoming mass treatment campaign against the disease.<br>- Carries out a headcount of household members and pre-marking of houses in his community.<br>- Detects and notifies suspected cases of yaws and other skin NTDs in his community as he does headcounts. |
|  | Azithromycin distributor | - Starts activity on the first day of the mass treatment campaign period and continues for the 3-days campaign period alongside the other two members of the team.<br>- Collects the daily quantity of AZC tablets required from the Health Area Chief<br>- Move door-to-door/ schools in his community and administer AZC to persons as from 6 months of age and above.<br>- Treats at least 200 persons per day.<br>- Notes and reports cases of refusals.<br>- Detects and notifies suspected cases of yaws and other skin NTDs in his community.<br>- Detects and notify cases of AZC side effects to proximity supervisor.<br>- Together with the registrar, makes an inventory of azithromycin used/lost or destroyed/ remaining at the end of each day.<br>- Returns remaining tablets of AZC to the Health Area Chief at the end of each day. |
|  | Registrar | - Starts activity on the first day of the mass treatment campaign period and continues for the 3-days campaign period alongside the other two members of the team.<br>- Move door-to-door/school alongside the distributor and tallies all persons |
|  |  | treated with AZC.<br>- Fills case-notification form of clinical cases of yaws and other skin-NTDs detected during period.<br>- Fills the pharmaco-vigilance form for any case of side effects and notify to the proximity supervisor.<br>- Together with the distributor, makes a daily synthesis of persons treated with AZC; cases of refusal; clinical cases of yaws and other skin-NTDs detected.<br>- Together with the distributor, transmit the daily summary data to the proximity supervisor latest 17h00 each day of the campaign. |
| Proximity supervisor |  | - Answerable to the Health Area Chief who doubles as the Health Area Supervisor for the TCT Campaign in his/her area.<br>- Responsible for supervision and follow-up of 3-5 TCT Treatment teams.<br>- Reinforces sensitisation in his catchment area.<br>- Detect stockouts of AZC and make refills.<br>- Manages cases of refusals.<br>- Manages minor cases of side effects and refers moderate-to severe case to the integrated health centre or district hospital.<br>- Cross-check the tally sheets, the pharmaco-vigilance form and the skin-NTD case-notification form for completeness and correctness.<br>- Verifies concordance between number of AZC tablets used and the number of persons treated by each team.<br>- Make daily syntheses of treatment data for all the teams under his supervision and transmit to the Chief of health area latest 17h30 each day of the campaign.<br>- Assist the Health Area Chief in the compilation of daily treatment data and in producing the daily synthesis for all the teams in the health area. |
| Health Area Chief / Health area supervisor |  | - Ensures the coordination of TCT campaign activities in his/her health area.<br>- Sensitizes community leaders and community members on the TCT Campaign in his/her area, while seeking their full support<br>- Designates and assigns TCT treatment teams.<br>- Defines itineraries for each TCT treatment team.<br>- Supply each TCT treatment team with azithromycin tablets, material, and tools.<br>- Coordinates proximity supervisors on the field.<br>- Manages cases of refusals<br>- Manages cases of minor to moderate side-effects and refer severe cases to the district hospital.<br>- Verifies concordance between number of AZC tablets used and the number of persons treated by each team.<br>- With the assistance of proximity supervisors, produces daily syntheses of treatment data for all the teams in his/her health area.<br>- Transmit the daily report of the health area to the health district latest 18h30 each day of the campaign.<br>- Makes an inventory of azithromycin used/lost or destroyed/ remaining at the end of each day.<br>- Returns remaining tablets of AZC to the Health district at the end of each TCT Campaign<br>- Ensures payment of actors of the TCT Campaign in his/her health area<br>- Produces a technical and financial report of the TCT Campaign in his/her health area.<br>- Transmit financial justification documents to the health district. |
| District supervisor to health area |  | - Ensure effectiveness of the TCT Campaign in the health area of assignment<br>- Assists the Health Area Chief in the implementation of the TCT Campaign<br>- Coordinates compilation and synthesis of daily TCT Campaign data.<br>- Transmit the daily report of the health area to the health district latest 18h30 each day of the campaign. |
| Health district supervision team | District Medical Officer (DMO) | - Participates in the national planning and evaluation meetings of the TCT campaign.<br>- Ensures the organization and coordination of TCT campaign activities in |
|  |  | his/her health district.<br>- Conduct advocacy with, local administrative, religious, and political leaders as well as other influential stake holders of the health district.<br>- Ensures community sensitization and mobilisation towards the TCT campaign.<br>- Organizes the management of rumours, resistance, and refusals.<br>- Ensure financial management of the TCT campaign.<br>- Prepares and transmit the technical and financial report of the TCT campaign for his/her health district.<br>- Ensures stock management of azithromycin and other supplies for the TCT campaign in his/her health district.<br>- Together with the regional supervisor assigned the district, co-presides daily debriefing meetings. |
|  | Chief of Bureau for Health (CBH) | - Assists the DMO in the coordination of TCT campaign activities.<br>- Participates in advocacy, community sensitization and mobilization towards TCT campaign.<br>- Participates in the management of rumours, resistances, and refusals.<br>- Ensures daily management of TCT campaign data, and transmission of daily syntheses to the regional and national coordination units latest 20h00 each day of the campaign period.<br>- Assists the DMO in the preparation of the technical and financial report of the TCT campaign.<br>- Ensures reception of the district stocks of azithromycin, redistribution to the health areas for the TCT campaign, and recollection of leftovers after the campaign. |
|  | District Communication Focal Person | - Coordinates under the supervision of the DMO, community sensitization and mobilization activities towards the TCT campaign.<br>- Supervises the social mobilizers in the pre-marking and household census.<br>- Participates in the management of rumours, resistances, and refusals.<br>- Produces a report of community sensitization and mobilization at the end of the TCT campaign. |
|  | District data manager | - Ensures collection of daily reports from the health areas<br>- Verifies timeliness and completeness of the reports from the health area.<br>- Interacts with the health area chiefs to ensure quality of the data received from them.<br>- Assists the Chief of Bureau for Health in the analyses and synthesis of the campaign data |
| National / Regional Supervisors |  | - Verifies the effectiveness and supervises the activities of TCT campaign in the health districts assigned.<br>- Supports the DMO in some technical tasks related to the TCT campaign.<br>- Supports the DMO in the management of rumours, resistances, and refusals.<br>- Provide solutions and give orientations to resolve challenges on the field.<br>- Ensures rapid supply of azithromycin to health areas reporting shortages.<br>- Ensure quality of the TCT campaign data and reports produced at the level of the health district.<br>- Collects and verify financial justification documents of the campaign from the DMO and transmit them to the national coordination unit.<br>- Produce a detailed supervision report. |
| Coordination |  | - Ensure strategic planning, organization, coordination, and supervision of the TCT campaign in all the districts involved.<br>- Ensure high level advocacy towards the campaign.<br>- Ensure effectiveness of the campaign in all health districts involved.<br>- Provide necessary logistics support for successful implementation of the TCT campaign in all |

|  |  |
| --- | --- |
|  | the health districts involved. |
|  | - |

Equally, the various tools required for the TCT campaign were adopted; the 3–4-day period for the campaign was fixed; the chronogram of activities for the campaign validated; the allocation of the number of teams, the required quantities of AZC 500mg tablets and other materials for each targeted health district was validated. Table 3 show a detail list of the tools developed and used for the TCT campaign in the Congo Basin of the CEMAC sub-region.

**Table 3:** List of tools developed/adapted for TCT campaign in the Congo Basin.

| N° | Tools | Tool Usage | Tool Holder |
| --- | --- | --- | --- |
| 1. | Chronogram of TCT Campaign activities | Indicates dates, periods, and venues of various campaign activities | All TCT campaign actors |
| 2. | Teams, AZC tablets and material allocation sheet | Indicates number of teams, AZC tablets and material allocated to each targeted district | Health District Management Teams, Supervisors and Coordination teams |
| 3. | Tally sheets | Score of people treated in the community during mass distribution of azithromycin | Mass AZC treatment teams |
| 4. | Azithromycin side effects monitoring forms | Recording of azithromycin side effects during mass distribution | Mass AZC treatment teams |
| 5. | Integrated skin-NTD case reporting form | Recording and notification of cases of skin NTDs detected | Mass AZC treatment teams, proximity supervisors |
| 6. | Review form | Registration of households to be revisited during mass distribution | Mass AZC treatment teams |
| 7. | Daily report form | Synthesis of daily treatment data for teams | Proximity supervisor |
| 8. | Daily summary form of the health area | Summary of daily mass distribution data for azithromycin | Health Area Supervisor |
| 9. | Daily summary form of the health district | Summary of daily mass distribution data for azithromycin | Health district Supervisor |
| 10. | Actors job description guide | Defines the role played by each actor in the implementation of TCT Campaign | All the actors |

The number of TCT-teams required for each health district was determined based on its population, such that a team was expected to treat 150 to 200 persons per day, and about 600 persons for the 3–4-day campaign period. The allocation of AZC 500mg tablets to health districts was estimated based on the population at a rate of 3 tablets per person on the average(25) (Table 4).

**Table 4.** Allocation of Azithromycin 500 mg tablets and teams by health district for the 1^st^ round of TCT.

| Health District | Total pop.<br>Targeted | N° of AZC<br>500mg tablets<br>allocated | TCT Campaign Teams |  |  |  |  |  |
| --- | --- | --- | --- | --- | --- | --- | --- | --- |
|  |  |  | Number of<br>TCT Teams | N° of<br>persons in<br>the TCT<br>Team | N° of<br>proximity<br>supervisors | N° if Health<br>Area<br>supervisors | N° of district<br>supervisors<br>to Health<br>Area | N° of<br>Regional /<br>National<br>supervisors |
| Cameroon |  |  |  |  |  |  |  |  |
| Abong-Mbang | 86,089 | 258,267 | 143 | 430 | 29 | 12 | 12 | 2 |
| Doume | 53,094 | 159,282 | 88 | 265 | 18 | 12 | 12 | 2 |
| Lomie | 37,668 | 113,004 | 63 | 188 | 13 | 8 | 8 | 2 |
| Mbang | 34,274 | 102,822 | 57 | 171 | 11 | 9 | 9 | 2 |
| Messamena | 30,023 | 90,069 | 50 | 150 | 10 | 10 | 10 | 2 |
| Moloundou | 48,077 | 144,231 | 80 | 240 | 16 | 8 | 8 | 2 |
| Ndelele | 101,680 | 305,040 | 169 | 508 | 34 | 10 | 10 | 2 |
| Nguelemendouka | 39,186 | 117,558 | 65 | 196 | 13 | 8 | 8 | 2 |
| Yokadouma | 118,598 | 355,794 | 198 | 593 | 40 | 13 | 13 | 2 |
| Djoum | 45,722 | 137,166 | 76 | 229 | 15 | 8 | 8 | 2 |
| Total Cameroon | 594,411 | 1,783,233 | 991 | 2,972 | 198 | 98 | 98 | 20 |
| Republic of Congo |  |  |  |  |  |  |  |  |
| Impfondo | 130,625 | 391,875 | 218 | 653 | 34 | 7 | 5 | 5 |
| Enyelle Betou | 109,643 | 328,929 | 183 | 548 | 37 | 4 | 4 | 2 |
| Ouessou | 96,525 | 289,575 | 161 | 483 | 31 | 7 | 7 | 5 |
| Sembe | 37,201 | 111,603 | 62 | 186 | 12 | 3 | 8 | 2 |
| Total Congo | 373,994 | 1,121,982 | 623 | 1,870 | 114 | 21 | 24 | 14 |
| Republic of<br>Central Africa |  |  |  |  |  |  |  |  |
| Boda | 122,516 | 367,548 | 204 | 613 | 51 | 12 | 4 | 12 |
| Mbaiki | 208,447 | 625,341 | 347 | 1,042 | 87 | 21 | 21 | 4 |
| Sangha-Mbaere | 149,976 | 449,928 | 250 | 750 | 62 | 16 | 8 | 8 |
| Total RCA | 480,939 | 1,442,817 | 802 | 2,405 | 200 | 49 | 33 | 24 |
| Grand Total | 1,449,344 | 4,348,032 | 2,416 | 7,247 | 513 | 168 | 155 | 58 |

The shipment of azithromycin and other material to the various TCT implementing health districts in Cameroon was done in collaboration with the Regional Fund for Health Promotion (RFHP), a regional structure responsible for procurement and supply chain management of essential drugs to the health facilities in their respective region. In the RCA and The Republic of Congo, this was carried out directly by the respective national yaws control programmes.

#### 1.4.2. High level advocacy

The second key activity of the planning phase was high level advocacy. This was conducted by a team comprising National Programme Manager, and Representatives of National support partners. The advocacy was targeted to the hierarchy of the Ministry of Public Health, and the administrative authorities (Governor & Prefects) of the regions and administrative divisions concerned by the TCT campaign. These advocacy meetings were used to explained to these authorities the stakes of the campaign, and to seek their full involvement in terms of administrative facilitation, community mobilisation and security guarantees necessary for a successful implementation of the campaign.

#### 1.4.3. Health district-level preparations

The third aspect of the planning phase were health district-level preparations, development of locally adapted sensitisation material, and community mobilization. District level preparations included: micro-planning, training of local supervisors and TCT campaign teams, reception and prepositioning of azithromycin and materials at the health area (sub-district) and community levels in readiness for the TCT campaign.

##### i. Community sensitisation and mobilization

Radio slots and jingles were developed in both the French and local languages spoken in each of the seventeen health districts involved in the TCT campaign (links to slots & jingles). These slots and jingles were broadcasted in all community radios located in the targeted health districts several times daily, and beginning at least one week to the launching of the campaign and throughout the 3-4-days period of the campaign. In some districts, call-in interactive radio programmes were held by the DHMTs, where they responded directly to questions from the community members regarding the TCT campaign. In addition to radio sensitization, messages from the district DHMTs were also read in churches and mosques calling on the population to massively turn out for the campaign.

##### ii. Pre-marking of households

Another community sensitisation and mobilisation strategy were the deployment of community mobilizers, who moved in the community from door-to-door passing on information on the campaign. During the door-to-door sensitisation, the mobilizers also carried out pre-marking of all the households in their respective communities. The pre-marking consisted in identifying to total number of persons living in each household and marking this number of the doorpost of the household. This number served as the denominator for the number of persons treated by the treatment teams when they reached the household during the actual TCT campaign period.

### 1.5. Execution of the 3-4-day TCT Campaign

#### 1.5.1. Official launching of the TCT Campaign

Each round of the TCT Campaign witnessed an official launching, by a high-level government official, in one of the health districts involved, in each of the three countries. The launching ceremonies were also carried out simultaneously in the remaining health districts by local administrative authorities. The ceremonies underscored the importance national governments attached to this health intervention as well as reinforced mobilisation of the population towards the campaign.

#### 1.5.2. Door-to-door treatment of the population

A 3-4-days of intensive treatment of the population with a single dose of azithromycin, immediately followed the official launching ceremonies of the TCT campaigns. Treatment teams of three persons each moved from door-to-door, in pigmy camps, quarters and villages according to pre-determined itineraries. In each household, the team mobilized the members following the data from pre-marking, treated each person present and who was 6 months or older with a single dose of azithromycin, coloured the left fifth finger of each treated person with an indelible marker, and indicated on the doorpost the number of persons treated. The number of persons treated was also tallied on the sheet designed for the purpose according to age and sex. The treatment team also identified and noted members of the household presenting with suspected lesions of skin NTDs including yaws, leprosy, Buruli ulcer. Any such persons were notified the supervisors who did clinical examination and conducted serological tests on the spot and/or collected samples when necessary.

#### 1.5.3. Supervision of the TCT Campaign

Four levels of supervision were organized for the TCT Campaign. Firstly, there were proximity supervisors who oversaw three to five treatment teams each. They worked under the supervision of the health area (sub-district) nurse, who played the role of the health area supervisor. The health area supervisor, who was assisted by a district supervisor assigned from the health district office worked under the direct supervision of District Medical Officer. The District Medical Officer was the main organizer of the TCT campaign in his/her health district. He was supervised by the regional and/or national level supervisor. Each of these levels of supervision had clearly defined job descriptions (Box 2). The aim of the supervision was to ensure that every activity of the campaign as well as data collection and reporting were carried out correctly as planned.

#### 1.5.4. Pharmacovigilance

The likelihood participants developing minor adverse events (AE) following azithromycin ingestion was low, and that of severe AE was very rare(26,42). Pharmacovigilance was however implemented during all of the campaigns to monitor for adverse events of the azithromycin being administered and to manage cases developing adverse effects during the campaign according the national pharmacovigilance guidelines(43). Prior to the TCT campaign, the treatment teams and the supervisors were trained on the identification and management procedures of minor, moderate and severe adverse events of azithromycin. They were then provided with pharmacovigilance forms for reporting on the adverse events. Cases with adverse events were managed.

##### Data production, collection, and management

During the TCT, data was produced on community treatment, azithromycin stock management, pharmacovigilance and detection of skin NTDs. The treatment teams used tally-sheets to record treatment data including number of persons treated per community by age and sex, the number of azithromycin tablets used, lost, and returned. They also used other specific forms to record suspected cases of skin NTDs, and adverse effects of azithromycin.

At the end of each day the proximity supervisors together with the teams under their supervision, made a synthesis of the treatment data from the tally-sheets using a daily report form, as well as synthesis of cases of skin-NTDs detected, synthesis of cases of adverse drug effects, and transmitted to the health area supervisor.

The health area supervisor assisted by the district supervisor to the health area, in turn, compiled and synthesized the reports from the teams through the proximity-supervisor, using the daily summary form of the health area, and transmitted to the district health management team at the end of each campaign day. At the level of the health district, the daily data from the health areas were compiled and synthesized using templates designed on Microsoft Excel spreadsheets. Feedback on the data were made by the district team to the health areas, requesting for clarification and corrections when necessary. The daily data from the district was then transmitted to national coordination team by email or other media platforms.

The data manager at the national coordination team compiled and synthesized data from all the health districts daily. These data were used to monitor the daily coverages by health district. Immediate feedback and orientations were given to the districts by the national coordination team, so that corrective measures were implemented for improvement of coverages and handling of major issues that could influence the TCT negatively.

### 1.6. Evaluation of the TCT Campaign

Evaluation of each round of the TCT was done at the health district and at the national levels.

#### 1.6.1. Evaluation of the TCT campaign at the health district

The evaluation of the campaign at the health district level was carried out within three days after the end of the 3-4-day TCT campaign period. The evaluation meeting presided at by the District Medical Officer, that brought together, the health area supervisors, the district supervisors, and the health district management team. The aim was to finalize the compilation and verification the campaign data from all the health areas, cross-check the coherence between number of persons treated and the amount of azithromycin tablets used, confirm the quantities of AZC tablets returned, before validating the data. Secondly, it was to assess the performance of each health area especially in terms of geographic and therapeutic coverages and deliberate on the challenges encountered and the possible solutions to them. The health district level evaluation was sanctioned by a final report of the campaign that was transmitted to the national coordination.

#### 1.6.2. The national level evaluation of the TCT campaign

The evaluation of each round of TCT at the national level was conducted within one week of the end the campaign. Presided over by the National Programme Manager, the evaluation, brought together national level stakeholders from the national programme, the WHO country office and support partners, the Regional Delegation of Public Health and the District Health Management Teams of the implementing health districts. In these evaluation meetings data and reports related to the TCT campaign were scrutinized, and then validated district by district. The validated data were then synthesized to produce the national level data, that was then published as the final campaign results. Challenges from the field were discussed and recommendations formulated for improvements.

### 1.7. Ethical considerations

All participants or their parents or guardians gave verbal informed consent before there were screened and treated with oral azithromycin. In addition, written informed consent was obtained from persons presenting with yaws-like skin lesions or from their parents or guardians for children who had lesions before finger-pricks blood sample was collected for on-the-spot serology testing and/or swabs were collected for PCR analyses at the national reference laboratory. The intervention was approved by the Cameroon National Ethics Committee for Research in Human Health (Ref. N° 2020/12/1327/CE/CNERSH/SP), and the Ethics Committee for Research in Health Sciences of the Republic of Congo (N° 0060/MESRSIT/IRSSA-CERSSA). Administrative authorisations were also obtained from the ministries of health in the three countries involved in the intervention. The supporting partners of the intervention had no influence on the design, data collection, analysis and interpretation, nor in the writing of the manuscript.

## 2. RESULTS

### 2.1. Therapeutic coverage

Table 4 gives detail of therapeutic coverages of the first round of TCT by country and health district. Out of the 1,530,014 persons targeted for the first-round TCT with azithromycin against yaws in 17 health districts of the Congo Basin, 1,456,691 persons were treated for an overall therapeutic coverage of 95.21%. The therapeutic coverages varied from 92.92% in Cameroon through 96.21% in the Republic of Congo to 96.96% in the Republic of Central Africa.

**Table 4.** Therapeutic Coverage of first round TCT for the three countries.

| Countries/ Health Districts | Target Population | N° of persons Treated | Therapeutic coverage | 95%CI for Therapeutic coverage |  |
| --- | --- | --- | --- | --- | --- |
|  |  |  |  | Lower | Upper |
| **Overall coverage for the three countries** | **1,530,014** | **1,456,691** | **95.21%** | **95.17%** | **95.24%** |
| **Cameroon** | **594,411** | **552,356** | **92.92%** | **92.86%** | **92.99%** |
| Abong Mbang | 86,089 | 77,330 | 89.83% |  |  |
| Doume | 53,094 | 44,089 | 83.04% |  |  |
| Lomie | 37,668 | 33,505 | 88.95% |  |  |
| Mbang | 34,274 | 32,041 | 93.48% |  |  |
| Messamena | 30,023 | 32,203 | 107.26% |  |  |
| Moloundou | 48,077 | 52,234 | 108.65% |  |  |
| Ndelele | 101,680 | 90,930 | 89.43% |  |  |
| Nguelemendouka | 39,186 | 35,620 | 90.90% |  |  |
| Yokadouma | 118,598 | 109,768 | 92.55% |  |  |
| Djoum | 45,722 | 44,636 | 97.62% |  |  |
| **Republic of Congo** | **373,994** | **359,810** | **96.21%** | **96.15%** | **96.27%** |
| Impfondo | 130,625 | 123,740 | 94.73% |  |  |
| Enyelle Betou | 109,643 | 115,108 | 104.98% |  |  |
| Ouessou | 96,525 | 87,474 | 90.62% |  |  |
| Sembe | 37,201 | 33,488 | 90.02% |  |  |
| **Central African Republic (CAR)** | **561,609** | **544,526** | **96.96%** | **96.91%** | **97.00%** |
| Boda | 122,516 | 115,793 | 94.51% |  |  |
| Mbaïki | 289,117 | 272,105 | 94.12% |  |  |
| Sangha-Mbaere | 149,976 | 156,628 | 104.44% |  |  |

The second round of the TCT was implemented only in Cameroon. Fig. 3 below shows the therapeutic coverages by health district of the second round compared to that of the 1^st^ round of TCT in Cameroon. For the second round, 615,503 (95.73%; 95%CI: 95.68%-95.78%) people were treated out of a population of 642,947 targeted in the ten health districts. There was a 3-percentage-point increase in the therapeutic coverage in the second round compared to 92.92% in the first round (P-value<0.001).

**Fig. 3.**
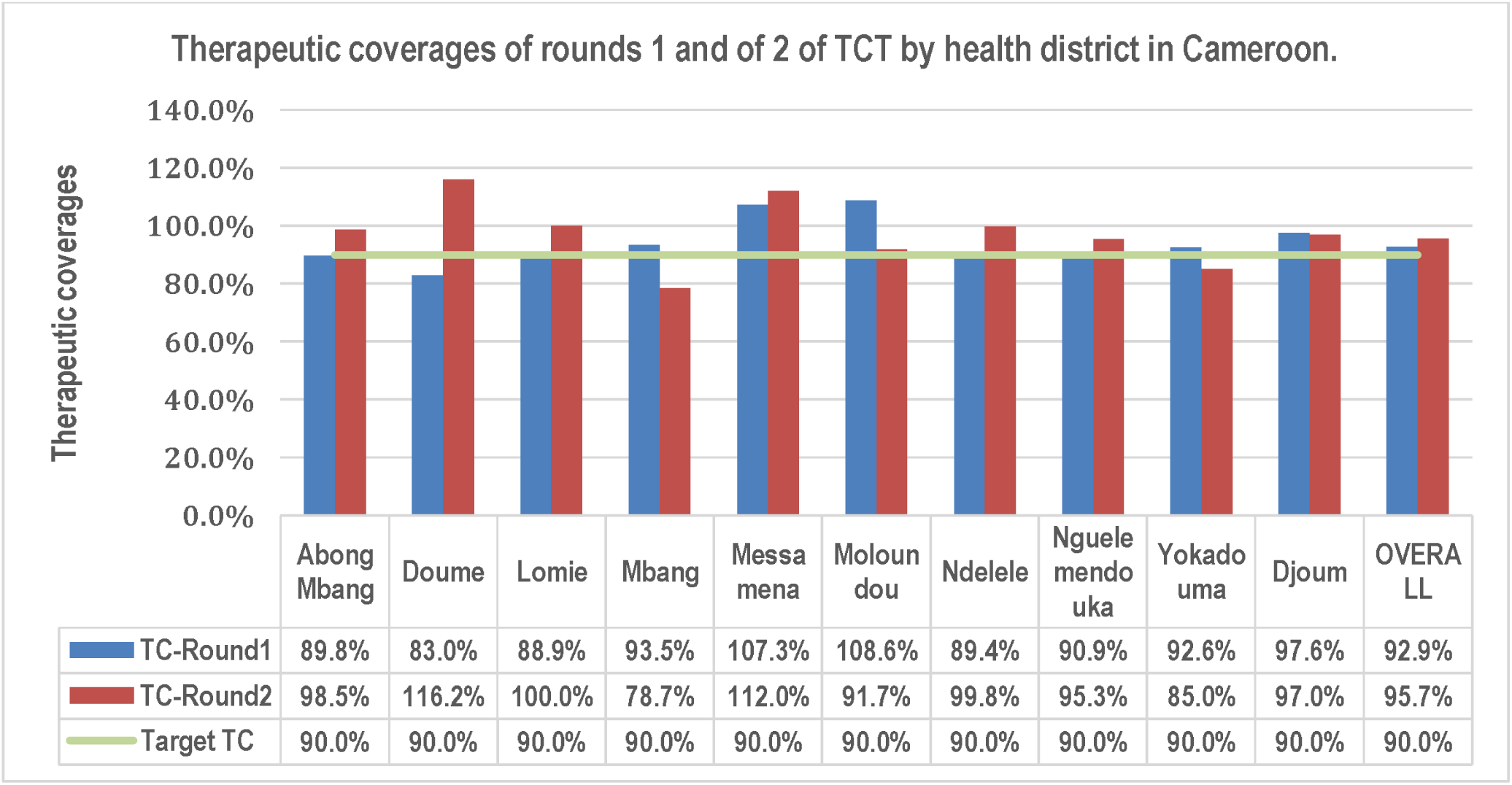
Therapeutic coverages (TC) of the 2^nd^ compared to the 1^st^ round of TCT in Cameroon. The TC of the second round was 3%-points over that of the first round. In both rounds. The TC were above the 90% target recommended by WHO.

Out of the ten health districts targeted for TCT in Cameroon, 9 of them harbour the Baka pigmy population. In the first round of TCT, the overall therapeutic coverage attained in this special population in Cameroon was 89.2%, slightly less than that the 90% recommended target. Efforts made during the second round, improved upon the therapeutic coverage to 93.1% in this population (P-value <0.001)(see Table 5).

**Table 5:** Therapeutic coverages of the Baka pigmy population in Cameroon.

| Health District | 1<sup>st</sup> Round TCT |  |  | 2<sup>nd</sup> Round TCT |  |  | P-value |
| --- | --- | --- | --- | --- | --- | --- | --- |
|  | Target Baka Pop | Pop Baka treated | Therapeutic coverage | Baka pop. Targeted | Baka pop. Treated | Therapeutic coverage |  |
| Abong Mbang | 5,405 | 4,131 | 76.4% | 5,676 | 6,690 | 117.9% |  |
| Djoum | 4,810 | 4,584 | 95.3% | 4,884 | 5,534 | 113.3% |  |
| Doume | 1,990 | 2,034 | 102.2% | 1,699 | 1,972 | 116.1% |  |
| Lomie | 12,754 | 11,431 | 89.6% | 12,849 | 10,304 | 80.2% |  |
| Mbang | 5,336 | 4,043 | 75.8% | 5,684 | 4,434 | 78.0% |  |
| Messamena | 2,924 | 2,895 | 99.0% | 2,853 | 2,866 | 100.5% |  |
| Moloundou | 6,056 | 6,069 | 100.2% | 6,443 | 6,096 | 94.6% |  |
| Ndelele | 2,677 | 2,114 | 79.0% | 2,676 | 2,324 | 86.8% |  |
| Yokadouma | 15,762 | 14,197 | 90.1% | 14,611 | 13,212 | 90.4% |  |
| **OVERALL** | **57,714** | **51,498** | **89.2%** | **57,375** | **53,432** | **93.1%** | **<0.001** |

### 2.2. Pharmacovigilance

During the TCT campaign in Cameroon, districts teams were required to detect and notify adverse effects of azithromycin treatment. This was more effective in two health districts compared to the other eight involved in the campaign (Table 6).

**Table 6.**
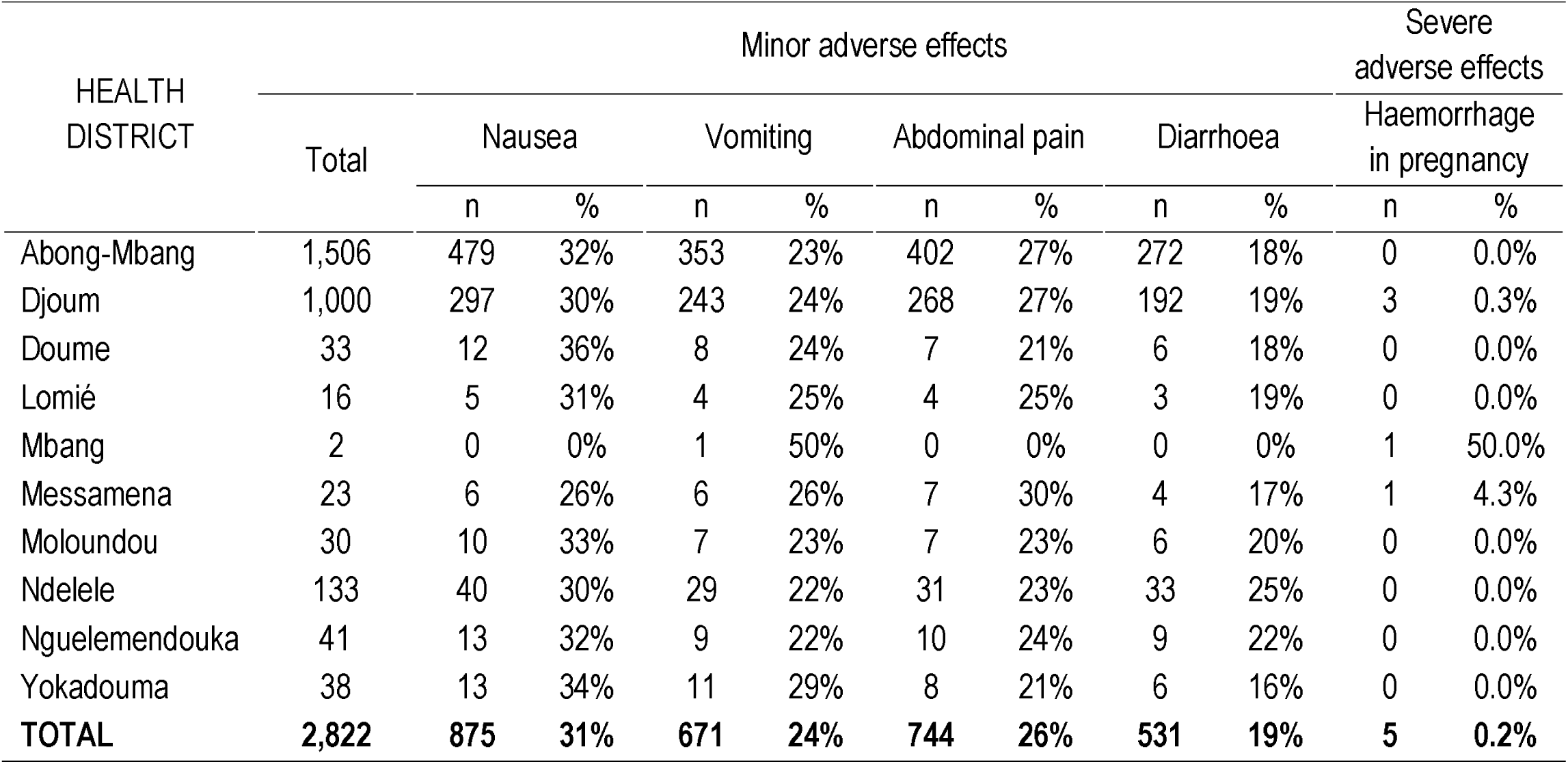
Adverse effects of azithromycin.

| HEALTH DISTRICT | Minor adverse effects |  |  |  |  |  |  |  |  | Severe adverse effects |  |
| --- | --- | --- | --- | --- | --- | --- | --- | --- | --- | --- | --- |
|  | Total | Nausea |  | Vomiting |  | Abdominal pain |  | Diarrhoea |  | Haemorrhage in pregnancy |  |
|  |  | n | % | n | % | n | % | n | % | n | % |
| Abong-Mbang | 1,506 | 479 | 32% | 353 | 23% | 402 | 27% | 272 | 18% | 0 | 0.0% |
| Djoum | 1,000 | 297 | 30% | 243 | 24% | 268 | 27% | 192 | 19% | 3 | 0.3% |
| Doume | 33 | 12 | 36% | 8 | 24% | 7 | 21% | 6 | 18% | 0 | 0.0% |
| Lomié | 16 | 5 | 31% | 4 | 25% | 4 | 25% | 3 | 19% | 0 | 0.0% |
| Mbang | 2 | 0 | 0% | 1 | 50% | 0 | 0% | 0 | 0% | 1 | 50.0% |
| Messamena | 23 | 6 | 26% | 6 | 26% | 7 | 30% | 4 | 17% | 1 | 4.3% |
| Moloundou | 30 | 10 | 33% | 7 | 23% | 7 | 23% | 6 | 20% | 0 | 0.0% |
| Ndelele | 133 | 40 | 30% | 29 | 22% | 31 | 23% | 33 | 25% | 0 | 0.0% |
| Nguelemendouka | 41 | 13 | 32% | 9 | 22% | 10 | 24% | 9 | 22% | 0 | 0.0% |
| Yokadouma | 38 | 13 | 34% | 11 | 29% | 8 | 21% | 6 | 16% | 0 | 0.0% |
| TOTAL | 2,822 | 875 | 31% | 671 | 24% | 744 | 26% | 531 | 19% | 5 | 0.2% |

Overall, 2 822 cases of adverse effects were notified. Over 99% were minor adverse effects including 875(31%) of nausea, 671(24%) of vomiting, 774(26%) of abdominal pains and 531(19%) of diarrhoea. Five (0.2%) cases of severe adverse effects notably haemorrhage in pregnancy were reported.

## 3. The impact of TCT on yaws prevalence

The third component in the implementation of the Morges strategy for yaws eradication is post TCT surveillance of yaws(22). Through the post TCT surveillance, the impact of the mass treatment campaign with azithromycin against yaws in the Congo Basin was evaluated. Table 7 show changes in the prevalences of clinical and DPP-serology confirmed yaws before and after TCT in three countries involved. During the post-TCT surveillance 22,286 persons were screened in the three countries. Of these, 1,421(6.4%; 95%CI: 6.1% -6.7%) clinical cases of yaws were detected. Serological testing with DPP syphilis screen and confirm serological test(41), revealed 83(0.37%; 95%CI: 0.30% - 0.46%) positive cases. With reference to the situation before TCT, there was a reduction in the prevalence of clinical yaws from 38.0% to 6.4% (P-value<0.001, OR=8.99; 95%CI: 8.35-9,67), and in the prevalence of DPP-serology confirmed yaws from 6.5% to 0.4% (P-value<0.001, OR=18.72; 95%CI: 14.75 – 24.01) (Fig.4A). There were however variations by country. In Cameroon, where 20,496 persons were screened in the ten health districts during the post TCT surveillance, 908 (4.43%; 95%CI: 4.15% - 4.72%) cases of clinically yaws were detected, of whom 48 (0.23%; 95%CI: 0.17% - 0.31%) were confirmed with DPP as cases of active yaws (Table 7). The prevalence of clinical yaws witnessed a drop from 29.9% to 4.4% (P<0.001, OR=9.21; 95%CI: 8.34 – 10.16) meanwhile that of serologically confirmed active yaws decreased from 6.31% to 0.23% (P<0.001, OR=28.67; 95%CI: 20.86 – 40.09) (Fig. 4B). In the Central African Republic, the prevalence of clinical yaws dropped from 32.5% to 29,0% (P-value = 0.063, OR=1.18 ; 95%CI: 0.99 – 1.40) meanwhile the prevalence of DPP confirmed yaws dropped from 2.4% to 0.8% (P-value=0.002, OR= 3.19; 95%CI:1.45 – 8.00) (Fig. 4C). In the Republic of Congo, the prevalence of clinical yaws decreased from 60.7% to 28.2% (P-value<0.001, OR=3.93; 95%CI: 3.24 – 4.78) and the prevalence of DPP confirmed yaws dropped from10.8% to 3.7% (P-value <0.001, OR=3.18; 95%CI: 2.09 – 5.01) (Fig 4D).

**Fig. 4.**
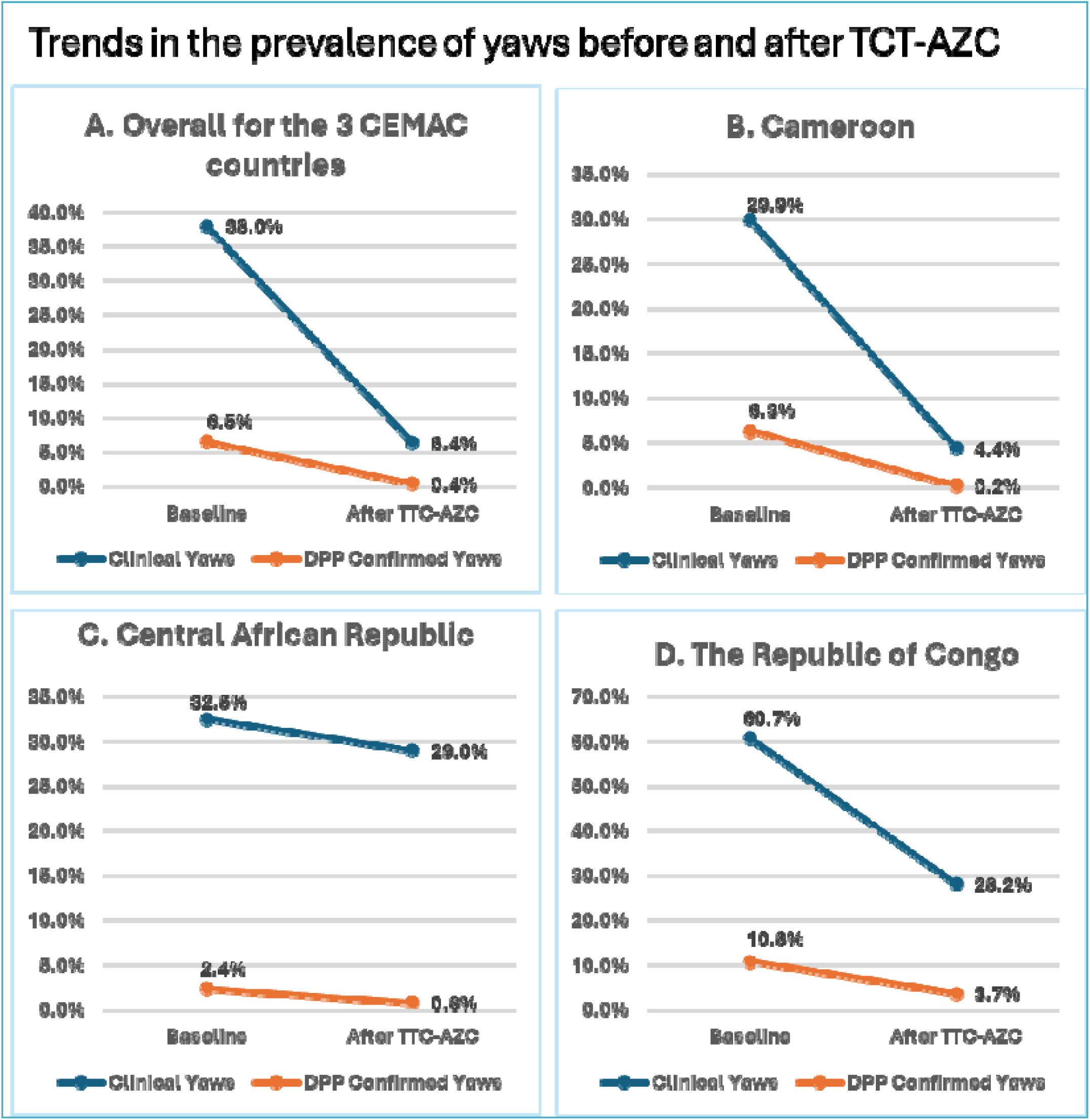
Trend in yaws prevalence at baseline and post-TCT in the 3 Congo-Basin countries. The overall prevalence of clinical yaws (Fig 4A) dropped from 38.0% to 6.4% meanwhile that of DPP confirmed (active yaws) dropped from 6.5% to 0.4%. For Cameroon, the prevalence of clinical yaws dropped from 29.9% to 4.4% and the DPP confirmed yaws dropped from 6.3% to 0.23%. (Fig. 4B). For CAR, the prevalence of clinical yaws dropped from 32.5% to 29.0% meanwhile that of DPP confirmed yaws dropped from 2.4% to 0.8% (Fig. 4C). In the Republic of Congo, the prevalence of clinical yaws reduced from 60.7% to 28.2% and that of DPP confirmed yaws dropped from 10.8% to 3.7% (Fig. 4D).

**Table 7.**
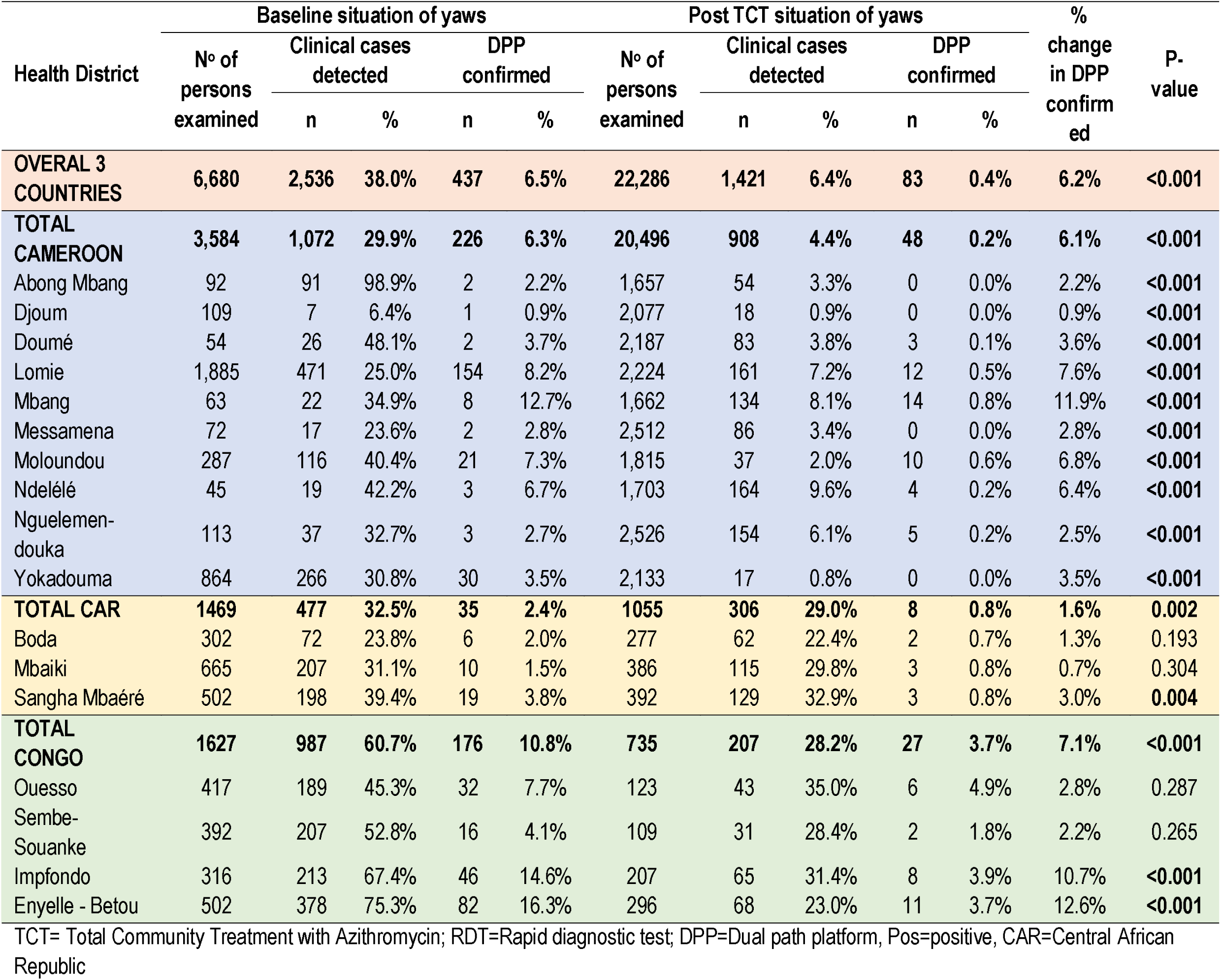
comparison of yaws prevalence at baseline and post TCT.

Post TCT surveillance show the presence of DPP-serologically confirmed yaws in 13 of the 17 health districts, including 6 out of 10 health districts in Cameroon and all 3 and 4 health districts in CAR and The Republic of Congo respectively. However, the prevalence had decreased by 6.2 percentage points, from 6.5% to 0.4% (P-value <0.001) overall in the three countries (Table 7 and Fig.4)

Fig. 4. show trends in the prevalence of yaws before and after TCT. In Cameroon, the prevalence rate of clinical yaws had declined from 29.9% to 4.43% (P-value <0.001, OR 9.21, 95%CI (8.34-10.16), meanwhile the prevalence rate of DPP-serology positive yaws dropped from 6.31% to 0.23% (P-value<0.001, OR=28.67, 95%CI (20.86-40.09) (Fig. 4B). In CAR, the prevalence of clinical yaws dropped slightly from 32.5% to 29.0% (P-value=0.063, OR= 1.18, 95%CI: 0.99-1.40) while that of DPP-serology confirmed yaws decreased from 2.4% to 0.8% (P-value=0.002, OR=3.19, 95% CI: 1.45-8.00) (Fig. 4C). In Congo, the prevalence of clinical yaws decreased from 60.7% to 28.2%, (P-value<0.001, OR=3.93, 95%CI: 3.24-4.78) meanwhile that of DPP-serology confirmed yaws dropped from 10.8% to 3.7% (P-value<0.001, OR=3.18, 95%CI: 2.09 – 5.01) (Fig. 4D).

The maps in Fig.5 further exemplify this reduction in yaws prevalence in the countries following TCT. Fig 5A and 5B shows the situation of clinical yaws in the 17 health districts that implemented TCT in the Congo Basin at baseline and after the mass treatment campaign. Fig. 5C and 5D, likewise, shows the evolution in the prevalence of DPP-serology confirmed (active) yaws in the same districts before and after the mass azithromycin treatment.

**Fig. 5.**
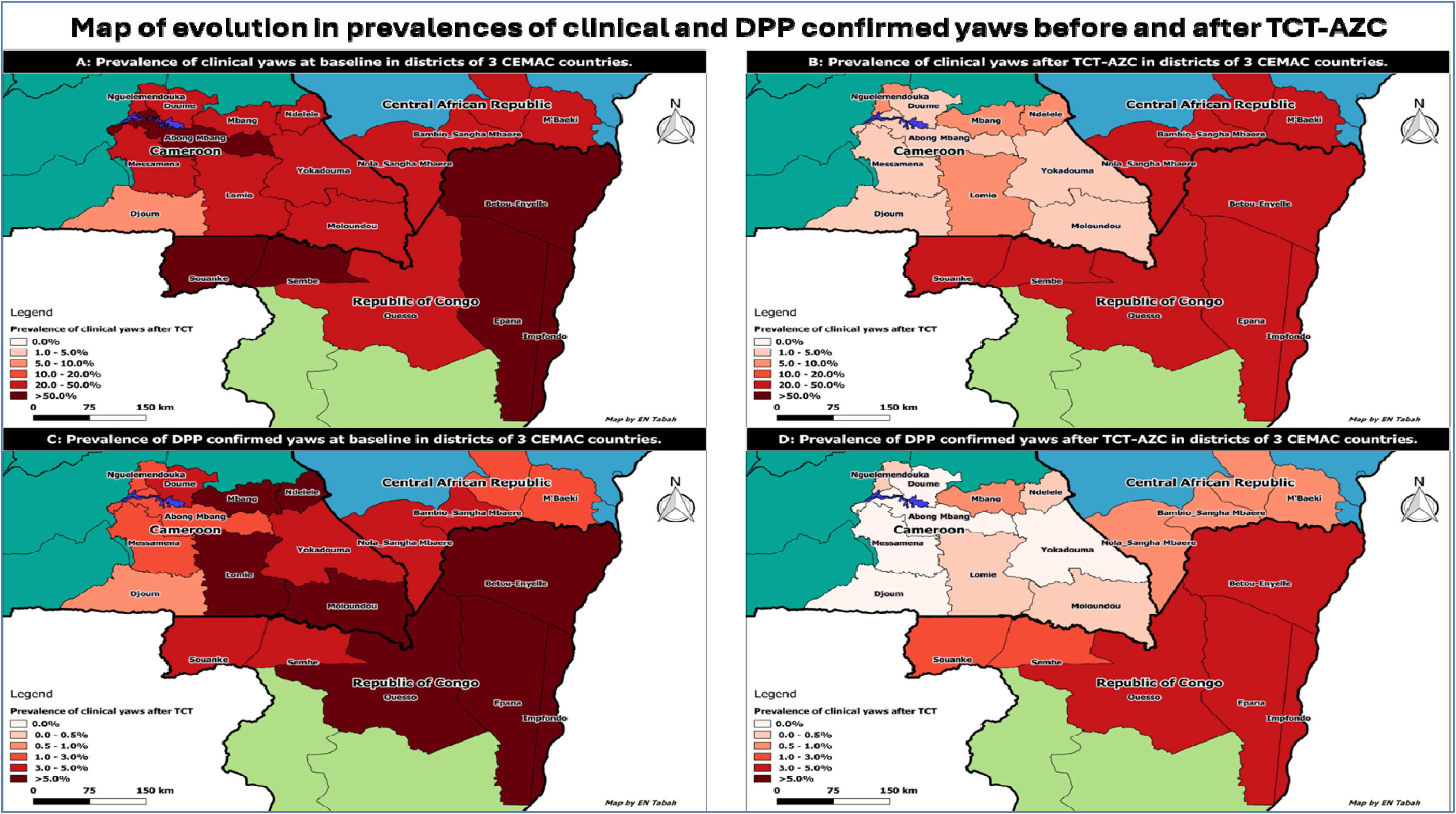
The prevalence of clinical yaws before (A) and after (B), and the prevalence of DPP confirmed yaws before (C) and after (D)TCT. The data for the situation after TCT is available only for oon, where we notice a remarkable drop in prevalence of clinical yaws to below 10% in all the 10 health districts as well as a remarkable drop in prevalence DPP confirmed yaws to 0% in h districts and to below 1% in the remaining 6 health districts targeted in Cameroon after the TCT.

## DISCUSSIONS

The renewed momentum for yaws eradication was manifested by the convening of the WHO Expert Consultative Committee in Morges, Switzerland from the 5^th^ to the 7^th^ of March 2012, during which the Morges Strategy for the eradication of yaws was developed(22,23). This was closely followed by the setting of a milestone for global yaws eradication by 2020, in the first global roadmap for overcoming the impact of NTDs for the period 2012 – 2020 (20) that has now been extended to 2030 in the second global roadmap for ending the neglect of NTDs to attain the sustainable development goals, published in 2021(44). One of the major components of the yaws eradication strategy is the mass drug administration with azithromycin, code-named Total Community Treatment with Azithromycin (TCT)(45), in serologically confirmed yaws endemic health districts.

In this report, we presented the first largescale implementation of TCT since the advent of the Morges strategy in 2012. The TCT implemented in the Congo Basin, was the largest in scale so far, given the size of the population covered of 1,530,014 people and the geographical extent of 17 health districts involved, spanning three countries of the CEMAC subregion namely Cameroon, The Central African Republic, and The Republic of Congo (Fig. 1). The scale of coverage is by far larger than those previously reported in the Lihir Island in Papua New Guinee where 16,092 persons were targeted(26), and in Abamkron sub-district in Ghana where 16,287 persons were targeted(27).

Prior to implementing TCT, serological surveys were carried out in the 17 health districts targeted to ascertain their yaws endemicity status as well as the level of prevalence (Table 2). The prevalence of active yaws varied from 17.83% in the Republic of Congo through 7.33% in the Republic of Central Africa to 6.31% in Cameroon. These served as baseline values against which the effect of TCT was going to be compared.

The TCT implemented in the CEMAC subregion, used a novel and unique model (Fig. 2) that was developed and successfully used in Cameroon, and later, in the Central African Republic and in The Republic of Congo respectively. The 3-component model included the planning, the execution, and the evaluation components; well-orchestrated in close successions, such that the whole process was implemented within a period of 15 to 21 days. The identification of key actors of the TCT campaign and the establishment of roles and job descriptions (Box 2) for them, was another important element in our model. Each actor used this as a guide for action regarding their defined roles.

The uniqueness of our model of TCT lies in: the 3-4-day campaign mode; the door-to-door community mass treatment with azithromycin; and the high level of community participation. The 3-4-day campaign duration is so far the shortest reported for TCT, compared to the two months period by *Médecins Sans Frontières* in Congo(46), and the relatively longer periods in Lihir(26), Papua New Guinea and in Ghana(27) respectively. The 3-4-day campaign period was charged with total effervescence in the communities, where every stratum beginning from the administrative, elected, religious, traditional authorities and the masses, were all mobilized towards the TCT. Community engagement and participation in the TCT was further marked by the implication of community volunteers in mass treatment teams in their own communities. Engaging 7,247 community volunteers was important to cover the 1,530,014-target population (Table 3) in three to four days, combing the hard-to-reach communities and pigmy camps embedded in the equatorial-rain forest of the Congo Basin on foot. These activities were meticulously coordinated and supervised by the local, regional, and national health authorities (Table 3). All these activities were essential for the successful implementation of the model.

The implementation of the model in varied local contexts and realities of the three countries of the CEMAC subregion, yielded very high therapeutic coverages above the 90% target recommended by the WHO(8) in all of these countries. All the three countries were involved in the first round of the TCT and registered an overall therapeutic coverage of 95.21%. By country, the therapeutic coverages varied from 92.92% in Cameroon through 96.21% in the Republic of Congo to 96.96% in the Republic of Central Africa. These coverage rates all stand above the 89% coverage registered in the Ghana(27) and the 82.7% coverage registered in Lihir, Papua New Guinea(26). For the second round of TCT conducted in Cameroon, the therapeutic coverage improved to 95.7% from the 92.92% registered in the first round, P<0.001, OR = 1.7 (1.68-1.73) (Fig.3). The therapeutic coverage among the most hard-to-reach indigenous Pigmy population also witnessed and increase from 89.2% in the first round to 93.1% in the second round, P<0.001, OR=1.64(1.57-1.71) (Table 5). This was an important achievement given that this population is the most vulnerable to yaws. In fact the resurgence of yaws in the Congo-Basin was among this population(40,46). With the high performance of our model, in different contexts, we would not hesitate to recommend it for use in other countries of Africa and beyond.

A pharmacovigilance system was put in place especially in Cameroon, that permitted the detection and notification of 2822 (0.5%) cases of adverse effects of azithromycin among the 594 430 people treated in the first round and 746 (0.12%) adverse events among the 615 503 people treated in the second round of TCT. The findings in Cameroon were comparable to 0.3% obtained in Ghana(27) but far lesser than the 17% reported in Lihir(26). Over 99% of the adverse effects notified in Cameroon were minor, and like in Lihir, included mostly nausea, vomiting, abdominal pains and diarrhoea. The low rate of adverse event reporting in our intervention could be explained by the lack of spontaneity in data reporting. This resulted probably from a common community belief “that it was natural for the people to have slight side effects after taking AZC tablets especially when the medicine had found a disease in the body and is combating it”.

The post-TCT surveillance enabled us to evaluate the impact of the mass treatment campaign with azithromycin against yaws. The TCT had a remarkable effect in the reduction of yaws prevalence in the CEMAC subregion, following the implementation of two rounds TCT campaign in Cameroon, and one round each in CAR and the Republic of Congo respectively. Overall, there was a nine-folds reduction in the prevalence of active yaws from 6.5% to 0.4% (P-value <0.001, OR=8.99, 95%CI: 8.35-9.67) in the three countries. The impact was greatest in Cameroon where a 28-folds reduction in the prevalence of active yaws was witnessed, compared to the 3-folds reduction seen in CAR and the Republic of Congo respectively. Just like was seen in 1956 in Nigeria(8), and in the pilots in the Lihir (26) and in Ghana(27), we have also shown that mass treatment with azithromycin, achieving therapeutic coverages of over 90% rapidly reduces the prevalence of yaws, and can potentially break the chain of its transmission.

The serological confirmation of active yaws cases in some of the health districts, suggests that two rounds of TCT with therapeutic coverages above 90%, are not sufficient to completely break the transmission chain. The question of whether conducting a third consecutive round of TCT would extinguish the transmission of yaws remains to be answered. While waiting, the reinforcement of surveillance and total targeted treatment (TTT) of confirmed cases of yaws, as recommended by WHO for low endemicity areas(25), should be the focus.

### Limitations

The major limitations we faces were contextual and included: the COVID-19 pandemic that slowed down field activities as well as the azithromycin procurement process. This particularly impacted the interval between TCT rounds, especially in Cameroon where the interval in 8 of the 10 health districts was extended from the recommended six months to 2 and half years. Secondly, we were operating in a difficult and hard-to-reach terrain of the equatorial rain forest of the Congo-Basin and targeting among others, the indigenous hard-to-reach Baka pigmy population. This had an impact on both the geographical and the therapeutic coverages in the first round of the TCT campaign. Thirdly, we were learning and doing at the same time, given that the novel model of the TCT campaign we used hard never been implemented elsewhere. So, the errors committed in the first round in Cameroon were corrected during the campaigns in CAR and the Republic of Congo as well as during the second round in Cameroon.

### Conclusions

We successfully implemented the first largescale TCT in the Congo Basin of the CEMAC subregion, using a novel model developed and employed in the local contexts of three countries namely Cameroon, CAR and the Republic of Congo. Five major lessons for a successful TCT were learnt in the process (Box 3). There was a remarkable reduction in the prevalence of active yaws in all the three countries following the TCT campaign especially in Cameroon that implemented two rounds. However, complete interruption of yaws transmission was not achieved even with two rounds of TCT. We recommend the TCT model we developed for use by any endemic country desiring to implement TCT. Secondly, we recommend at least three rounds of TCT per implementation unit for achievement of complete interruption of yaws transmission.

#### Box 3. Major lessons learnt for achieving a successful TCT

1. The use of a campaign strategy for Total Community Treatment with Azithromycin:

□ For a short duration of 3-4 days,
□ Using the door-to-door strategy,
□ Involving a large number of teams.
2. Meticulous planning and micro-planning at district and sub-district levels
3. Implication & Involvement of high-level administrative authorities
4. Community sensitization
5. Effective community mobilization and community participation.

## Authors contribution

ENT and AUB conceived and designed the intervention with inputs from KBA. ENT, AUB, SA designed the TCT model. ENT, AUB, DCA, GAA, BB, TNC, INN, LDP, BBE, KWCE, AS, GE, and ST implemented the intervention and gathered data and samples. ST, DV and SE were mostly responsible for analyses of biological samples. ENT did the statistical analyses. ENT, AUB, DCA, TNC, INN, LDP and BBE wrote the first draft of the report. All authors contributed to the revisions and approved the final version.

## Declaration of Interests

All the authors declare no competing interests.

## Data Availability

All relevant data are within the manuscript and its Supporting Information files.

## Acknowledgement

We acknowledge the village traditional rulers and populations of the intervention health districts for their adherence; the TCT teams, the health areas and the district health management teams for the efforts with the implementation; the local administrative authorities for their efforts in mobilizing the populations towards the intervention; the authorities of the Regional Delegations of Public Health for the East and South Regions of Cameroon, as well as the Sangha Mbaere and Lobaye Prefectures of the CAR; and the Likouala and Sangha Departments of the Republic of Congo for their supervision of the intervention on the field; the hierarchy in the ministries of health of the Cameroon, CAR and The Republic of Congo for guidance and general oversight. The implementation of the intervention was coordinated in each of the three countries by the National Programmes in charge of skin NTDs control. The intervention was funded through the Organisation for the Coordination of Epidemics Control in Central Africa (OCEAC). The FAIRMED provided administrative and technical support of the intervention in the three countries. The World Health Organisation in collaboration with EMS Brazil supported with azithromycin 500mg tablets; and the Centre Pasteur Yaoundé Reference Laboratory supported with analyses of biological samples.

## Financial Disclosure

The authors did not receive any funding for the writing of this manuscript.

## Notes

### Competing Interest Statement

The authors have declared no competing interest.

### Funding Statement

The author(s) received no specific funding for this work.

### Author Declarations

The intervention was approved by the Cameroon National Ethics Committee for Research in Human Health, and the Ethics Committee for Research in Health Sciences of the Republic of Congo. Administrative authorisations were also obtained from the ministries of health in the three countries involved in the intervention.

